# Prevalence of cirrhotic cardiomyopathy according to different diagnostic criteria. Alterations in ultrasonographic parameters of both left and right ventricles before and after stress

**DOI:** 10.1101/2022.11.02.22281851

**Authors:** Dimitrios S Karagiannakis, Katerina Stefanaki, George Anastasiadis, Theodoros Voulgaris, Jiannis Vlachogiannakos

**Author notes:** Corresponding author: Dimitrios S Karagiannakis. Academic Department of Gastroenterology, Medical School of National & Kapodistrian University of Athens, “Laiko” General Hospital, 17 Agiou Thoma street, 11527, Athens, Greece. All authors contributed to the interpretation of the data and reviewed and approved the manuscript.

## Abstract

**Background:** Prevalence of cirrhotic cardiomyopathy (CC) remains controversial. Several guidelines have been proposed for its assessment.

**Aim:** To estimate the frequency of CC by using all of the proposed diagnostic criteria, to describe the whole spectrum of cardiac alterations and investigate the role of stress in unmasking latent cases of CC.

**Methods:** Ninety consecutive patients were recruited. CC was evaluated by using the Montreal, the 2009 and 2019 criteria. Dobutamine stress test was also performed.

**Results:** LVDD was identified in 72(80%), 36(40%) and 10(11.1%) patients based on the above criteria, respectively. None of the patients had right ventricular systolic dysfunction, neither at rest, nor after stress. Stress test revealed left systolic dysfunction in 4(4.5%) patients. According to 2019 criteria, presence of LVDD was not associated with gender, etiology, or staging of liver disease. Patients with LVDD had longer QTc (p=0.002), larger LAvol (p=0.0001), lower TAPSE(s) (p=0.012), lower SRV(s) (p=0.0001) and lower ΔCI (p=0.009) compared to those without. Patients with Child-B/C had longer QTc (p=0.004), higher BNP (p=0.016), higher E/e’ (p=0.0001) and higher E/e’(s) (p=0.003), compared to Child-A patients. A significant correlation was demonstrated between Child-Pugh score and E/e’ (p=0.0001), or E/e’(s) (p=0.002).

**Conclusions:** In accordance with the recent guidelines the prevalence of CC seems to be lower. LVDD is the predominant feature of CC and aggravates along with the severity of liver disease. After dobutamine administration several sonographic variables exacerbate, particularly in Child-B/C patients, indicating a potential higher risk for clinical heart failure during stressful invasive interventions.

## Introduction

In cirrhosis, liver dysfunction and the presence of portal hypertension result in splanchnic arterial vasodilation due to overproduction, impaired degradation, and portosystemic shunting of vasodilator factors. The splanchnic arterial vasodilation and the reduced systemic vascular resistance leads to low blood pressure and reduced central blood volume with central or ‘‘effective’’ hypovolemia [1]. In order to compensate, the sympathetic nervous system is activated, leading to increased heart rate and output and to a hyperdynamic circulation. However, as liver dysfunction and portal hypertension are aggravating, the splanchnic vasodilation is worsening, making the increased cardiac rate and contractility unable to further counterbalance patients’ hemodynamic circulation. As a concequence, the renin-angiotensin-aldosterone axis is activated and vasopressin is released, in order to increase the blood pressure and the arterial blood volume [1]. Nonetheless, the hemodynamic state is remaining still extremely susceptible to factors that may influence the splanchnic arterial vasodilation, such as bacterial infections and overproduction of pro-inflammatory cytokines [2]. Furthermore, it seems that along with liver disease’s progression and portal pressure’s exacerbation, a derangement in cardiac function is also developed, leading to further arterial hypoperfusion and circulatory impairment [3]. This clinical entity is called “cirrhotic cardiomyopathy” (CC) and is characterized by altered diastolic relaxation, electrophysiological abnormalities and impaired contractility, under physiological or pharmacological stress, all occurring in the absence of other known causes of cardiac disease [4-6]. Diastolic dysfunction seems to precede, while systolic dysfunction is rarely present at rest, as the ejection fraction (EF%) usually is preserved, due to the arterial vasodilation and the concomitant reduced afterload. Any systolic abnormality is often unmasked under physiological or pharmacological stress [7]. Until now, the clinical significance of CC has been clarified only in cases of transjugular intrahepatic portosystemic shunt (TIPS) insertion, or liver transplantation [8,9], while its role on the prognosis of patients not undergoing any invasive procedure remains debatable [10-13]. Moreover, there is a disagreement amongst the researchers about the prevalence of CC, as different diagnostic criteria have been used for its evaluation in the studies published so far [14-16]. In 2016, the American Society of Echocardiography and the European Association of Cardiovascular Imaging proposed new guidelines for the diagnosis of left ventricular diastolic dysfunction (LVDD) in patients with normal EF% [17] and recently Izzy et al modified them in order to become more suitable for patients with cirrhosis [18]. The aim of this study is to evaluate the prevalence of CC according to all of the proposed guidelines, to underline the differences between them and to illustrate the ultrasonographic cardiac characteristics of cirrhotic patients according to the latest, modified criteria.

## Methods

### Patients

Over a period of 18 months, consecutive cirrhotic patients of any etiology and severity of liver disease, aged from 18 to 80 years old who attended our clinic, were considered eligible for inclusion into the study. The diagnosis of cirrhosis was based on clinical and laboratory findings, endoscopy and imaging studies and confirmed by liver elastography. Only patients with liver stiffness ≥13 kPa by two-dimension shear wave elastography (2D-SWE) were finally included [19]. Exclusion criteria were history of arterial hypertension, chronic cardiac, pulmonary or renal disease, diabetes mellitus, active bacterial infection, recent gastrointestinal bleeding (<1 month), hepatocellular carcinoma, recent or active ethanol abuse (<6 months) [20] and treatment with drugs that could affect cardiac function or circulatory parameters, like vasoactive drugs or nitrates. Active bacterial infection was ruled out by history, clinical examination, blood tests, culture of urine, chest radiograph, and in ascitic patients by culture and white cell count of ascitic fluid. Large-volume paracentesis was not performed in our ascitic patients during the last month before their recruitment in the study. Patients with alcoholic cirrhosis had prolonged periods of abstinence, confirmed by detailed history, discussion with relatives, and non-scheduled plasma alcohol determinations during their visits. Patients under treatment with beta-blockers for the prevention of variceal bleeding, had temporarily discontinued them at least 15 days prior to their cardiological assessment. The study protocol was approved by the Ethics Committee of General Hospital of Athens “Laiko”, Greece. A written consent was obtained from each patient with respect to all ethical guidelines issued by the 2000 revision (Edinburgh) of the 1975 Declaration of Helsinki.

### Electrocardiography and Echocardiography Protocol

Electrocardiograph (ECG) was recorded by a conventional electrocardiogram (Cardioline ar 600, Italy). QT intervals were corrected (QTc) with Bazett’s formula. A pulsed wave (PW) Doppler echocardiography with tissue Doppler imaging (TDI) (General Electric, Vivid 3, USA) was used to estimate the following cardiac parameters: heart rate (HR), left ventricular end diastolic diameter (LVEDD), left ventricular end systolic diameter (LVESD), left atrium volume (LAvol), left ventricular ejection fraction at rest (EF%), cardiac index (CI), systemic vascular resistance index (SVRI), peak early filling velocity during early ventricle diastole (E wave), late diastolic filling velocity during atrial systole (A wave), E/A ratio (E/A), deceleration time of E wave (DT), isovolumetric relaxation time (IVRT), early diastolic mitral annular velocity from the septal side (e’septal), early diastolic mitral annular velocity from the lateral side (e’lateral), average early diastolic mitral annular velocity (e’av), E/e’av. ratio (E/e’av.), pulmonary artery systolic pressure (PASP), tricuspid regurgitation velocity (TRV), tricuspid annular plane systolic excursion (TAPSE) and systolic right ventricular function (SRV). After the assessment of cardiac function at rest, a dobutamine stress test was performed and CI after stress [CI(s)], early diastolic mitral annular velocity from the septal side after stress [e’septal(s)], E/e’av after stress [E/e’av(s)], TAPSE after stress [TAPSE(s)], systolic right ventricular function after stress [SRV(s)] and tricuspid regurgitation velocity after stress [TRV(s)], were evaluated. Three long-axis and three short-axis slices (basal, mid-ventricular and apical) were acquired in order to cover 16 myocardial segments [21]. Serum brain natriuretic peptide (BNP) was estimated as well. Dobutamine was infused intravenously at 3-min stages. The initial dose was 2.5 μg/kg/min with gradual increase to 5, 7.5, 10, and 20 μg/kg/min and, if needed atropine injection with purpose to reach the maximum cardiac strain. Repeat short-axis images, as well as long-axis images, were acquired at the end of each stage. During dobutamine test, patient’s symptoms, heart rate, blood pressure, and electrocardiogram were monitored. All examinations were performed by a single, experienced cardiologist (GA). The results were stored digitally and analyzed offline twice in different periods of time. Differences were rarely found between the two measurements. If this happened, the average values were obtained.

### Criteria for the diagnosis of left ventricular diastolic dysfunction (LVDD)

a. Criteria of Montreal (2005) [22] E/A<1, or DT>200 msec, or IVRT> 80 msec
b. Criteria of the American Society of Echocardiography (2009) [23] e’ septal<8 cm/sec, e’ lateral<10 cm/sec, LAvol≥34 ml/m^2^
c. Criteria of the American Society of Echocardiography and the European Association of Cardiovascular Imaging (2016) [17], modified in 2019 by the CC consortium [18]. e’ septal<7 cm/sec, E/e’ septal≥15 cm/sec, LAvol>34 ml/m^2^, TRV>2.8 m/sec Three abnormal out of the 4 above parameters define LVDD. In case of 2 abnormal and 2 normal parameters, the LVDD cannot be assessed (indeterminate state), whereas if 1 parameter is normal and 3 abnormal, LVDD is excluded. Left ventricular diastolic function was further assessed after the performance of a dobutamine stress test, in order to uncover any latent diastolic dysfunction not apparent at rest.

### Criteria for the diagnosis of left ventricular systolic dysfunction (LVSD)

EF%≥55, or ≥50 at rest, according to Montreal or to more recent criteria respectively. An increase of EF% (ΔEF%), or CI (ΔCI) less than 10% at peak dobutamine infusion was also consistent with LVSD [17,22].

### Criteria for the diagnosis of right ventricular systolic dysfunction (RVSD)

TAPSE<17 mm, or SRV<9.5 cm/sec at rest, or TAPSE(s) <17 mm, or SRV(s) <9.5 cm/sec after dobutamine stress test [24,25].

### Statistical analysis

Statistical analysis was performed by using SPSS (SPSS software; SPSS Inc, Chicago, IL, USA). Quantitative variables were compared with independent student’s t-test or Mann-Whitney test for normally and non-normally distributed variables and data presented as mean ± SD, or median (range) respectively. Qualitative variables were compared with corrected Chi-squared test or two-sided Fisher’s exact test, as appropriate. The concordance of different diagnostic criteria was determined using the proportion of agreement, and inter-rater agreement kappa (k), which was interpreted as follows: <0.20 – poor, 0.21-0.40 – fair, 0.41-0.60 – moderate, 0.61-0.80 – good, 0.81-1.00 – very good. The relationship between parameters was estimated by using the Spearman’s correlation coefficient. All tests were two sided and p values <0.05 were considered to be significant.

## Results

### Demographic and Clinical Data

Over the study period, 107 cirrhotic patients visited our clinic. Seventeen of them were excluded, as three were diagnosed with liver cancer, five had known heart disease (coronary artery disease, valvular disorders etc), four had arterial hypertension and five were active drinkers. Therefore, 90 patients were finally recruited. Sixty out of 90 patients (66.7%) were males and 30 (33.3%) females. The median age was 55 (33-78) years. Patients’ demographics are summarized in table 1.

**Table 1.**
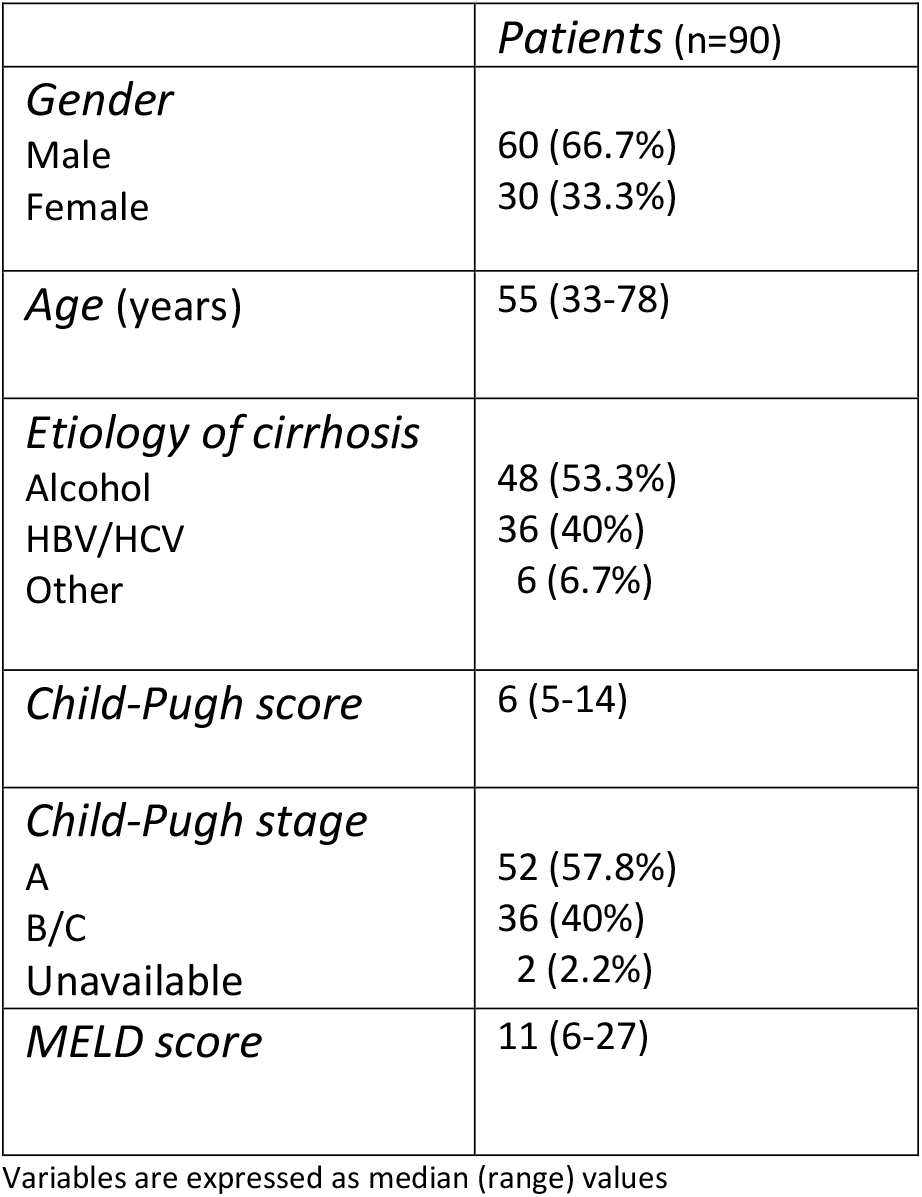
Patients’ characteristics

### Left ventricular diastolic dysfunction (LVDD)

According to Montreal criteria, 72 (80%) patients had LVDD. Using the criteria of 2009, 36 (40%) patients were diagnosed with LVDD. The agreement between the two methods was good (k value: 0.754; reclassification rate 11%, p<0.0001). Based on the latest criteria of 2019, 4/90 (4.45%) patients had LVDD, while 10/90 (11.1%) characterized as “indetermined”. The latter group of patients was further evaluated according to the proposed algorithm and 6 out of 10 were subsequently diagnosed with LVDD. Therefore, 10 out of 90 (11.1%) patients had LVDD in total. The percentage of patients detected with LVDD did not change after the administration of dobutamine, as no any new cases of LVDD distinguished after the re-assessment of left ventricular diastolic function at the peak of the test. The agreement between the latest criteria of 2019 and those of Montreal was fair (k value: 0.338; reclassification rate: 23%, p<0.001), while the agreement between the criteria of 2019 and those of 2009 was fair again (k value: 0.316; reclassification rate: 28.9%, p<0.0001).

The presence of LVDD according to Montreal criteria was not significantly associated with Child-Pugh stage (A vs B/C; p=0.21) or gender (p=0.575), while it had a trend towards the non-alcoholic etiology of liver disease (p=0.071). Presence of LVDD according to criteria of 2009 was significantly associated with male sex (χ^2^:4.583; p=0.032) and non-alcoholic etiology of liver disease (χ^2^:5.030; p=0.032), but not with Child-Pugh stage (p=0.575). LVDD development based on the recent guidelines was not significantly associated with gender (p=0.447), etiology of cirrhosis (alcoholic vs non-alcoholic) (p=0.505), or Child-Pugh stage (p=0.306).

### Left ventricular systolic dysfunction (LVSD) and right ventricular systolic dysfunction (RVSD)

Regardless of the criteria applied, LVSD wasn’t detected in our patients at rest, as no one had EF%<55%. Four patients (4.45%) were found incapable of increasing adequately the cardiac index (ΔCI<10%) during the dobutamine stress test. No RVSD was noticed at rest, as no one of the patients had TAPSE<17 cm/sec, or SRV<9.5 cm/sec. After stress test, no cases of RVSD were identified as TAPSE(s) and SRV(s) remained over 17 cm/sec and 9.5 cm/sec in all patients, respectively.

### Comparison of the ultrasonographic parameters between patients diagnosed with LVDD and those without, according to the latest guidelines

Patients with LVDD had significantly prolonged QTc [465(449-497) vs 433(368-492) msec;p=0.002], increased A [101(64-114) vs 76(42-127) cm/sec;p=0.007], lower E/A [0.59(056-1.4) vs 0.99(0.63-1.95);p=0.025], increased IVRT [120(118-127) vs 105(70-130) msec;p=0.0001], lower e’ septal [6.5(5-6.9) vs 8.85(5.3-15.3) cm/sec;p=0.0001], larger LAvol [38.3(24-42.9) vs 22.8(12.1-36.2) ml/m^2^;p=0.0001], higher TRV [2.9(2.5-3) vs 2.5(1.8-3) m/sec;p=0.009] and lower SVRI [1963(1517-2606) vs 2425.5(1355-3607) dyn/s/m^2^/cm^-5^;p=0.04], compared to patients without LVDD respectively. The former group of patients tended to be older in age (p=0.072), with higher BNP levels (p=0.081) and reduced e’lateral (p=0.064) compared to patients without LVDD, but the differences did not reach the statistical significance. Regarding of the dobutamine stress test parameters, patients with LVDD had statistically significant reduced e’septal(s) [8.3(7.5-12.7) vs 10.5(5.5-20.5) cm/sec;p=0.033], lower TAPSE(s) [27(23-28) vs 29(21-34) mm;p=0.012], lower SRV(s) [17.2(12.3-25) vs 26.3(16-41) cm/sec;p=0.0001], lower CI(s) [4(3.5-8) vs 5.2(2.4-10.5);p=0.022], lower ΔCI [0.3(0.13-0.86) vs 0.8(−0.04 – 2.52) L/min/m^2^;p=0.009] and lower ΔSRV [0.12(−0.07 – 0.34) vs 0.5(0.03-1.79) cm/sec;p=0.0001], in comparison to patients without LVDD respectively. The two groups had no differences regarding the Child-Pugh and the MELD score. Table 2 summarizes the differences between the two groups of patients.

**Table 2.** Differences between patients with or without LVDD

| Parameter | No LVDD | LVDD | p value |
| --- | --- | --- | --- |
| Age (years) | 55 (33-78) | 70 (45-77) | 0.072 |
| HR (bpm) | 76 (59-110) | 81 (59-95) | NS |
| QTc (msec) | 433 (368-492) | 465 (449-497) | 0.002 |
| EF (%) | 65 (56-75) | 64 (57-74) | NS |
| E (cm/sec) | 76.5 (50-128) | 64 (58-105) | NS |
| BNP (pg/ml) | 24.3 (5-286) | 106 (5-369) | 0.081 |
| A (cm/sec) | 76 (42-127) | 101 (64-114) | 0.007 |
| E/A (ratio) | 1 (0.6-2) | 0.6 (0.56-1.4) | 0.025 |
| DT (msec) | 220 (121-317) | 233.5 (220-253) | NS |
| IVRT (msec) | 105 (70-130) | 120 (118-127) | 0.0001 |
| TAPSE (mm) | 24 (21-37) | 23 (21-27) | NS |
| e' septal (cm/sec) | 8.9 (5.3-15.3) | 6.5 (5-6.9) | 0.0001 |
| e' lateral (cm/sec) | 12.4 (7.5-19.7) | 10.5 (7-16.5) | NS |
| E/e'av (ratio) | 7.2 (3.7-13.6) | 6.9 (6.6-10.6) | NS |
| SRV (cm/sec) | 16.5 (11.9-28) | 15.4 (13.2-18.7) | NS |
| CI (L/min/m<sup>2</sup>) | 3 (2.1-5.1) | 3.3 (2.4-4.3) | NS |
| LA (ml/m<sup>2</sup>) | 22.8 (12.1-36.2) | 38.3 (24-42.9) | 0.0001 |
| LVEDD (mm) | 50 (40-62) | 49 (42-62) | NS |
| LVESD (mm) | 31 (22-37) | 33 (24-37) | NS |
| PASP (mmHg) | 31.5 (18-43) | 35 (30-45) | NS |
| TRV (m/sec) | 2.5 (1.8-3) | 2.9 (2.5-3) | 0.009 |
| SVRI (dyn/s/m<sup>2</sup>/cm<sup>-5</sup>) | 2425.5 (1355-3607) | 1963 (1517-2606) | 0.04 |
| Child-Pugh score | 6 (5-14) | 7 (5-9) | NS |
| MELD score | 11 (6-27) | 11 (8-16) | NS |
| E(s) (cm/sec) | 76 (50-182) | 69 (49-137) | NS |
| e' septal(s) (cm/sec) | 10.5 (5.5-20.5) | 8.3 (7.5-12.7) | 0.033 |
| e' lateral(s) (cm/sec) | 13.5 (7.3-22) | 12.9 (9.3-21) | NS |
| E/e'av(s) (ratio) | 6.3 (3.3-10.2) | 6.9 (3.8-9.7) | NS |
| TAPSE(s) (mm) | 29 (21-34) | 27 (23-28) | 0.012 |
| SRV(s) (cm/sec) | 26.3 (16-41) | 17.2 (12.3-25) | 0.0001 |
| CI(s) (L/min/m<sup>2</sup>) | 5.2 (2.4-10.5) | 4 (3.5-8) | 0.022 |
| ΔSRV (cm/sec) | 0.5 (0.03-1.8) | 0.12 (-0.07 – 0.34) | 0.0001 |
| ΔTAPSE (mm) | 0.14 (-0.18 – 0.42) | 0.95 (0.04-0.17) | NS |
| ΔCI (L/min/m<sup>2</sup>) | 0.8 (-0.04 – 2.5) | 0.3 (0.13-0.86) | 0.009 |
| ΔE/e'av | -0.11 (-0.43 – 0.6) | -0.38 (-0.44 – 0.06) | NS |

Variables are expressed as median (range) values

### Differences in echocardiographic parameters between compensated and decompensated cirrhotic patients

Patients with Child-Pugh stage B/C had statistically longer QTc [454(416-497) vs 428(368-490) msec;p=0.004], increased E [90(51-128) vs 72(53-119) cm/sec;p=0.032], higher BNP levels [46(6.8-369) vs 18.5(5-145) pg/ml;p=0.016], increased A [87(53-127) vs 72(53-119) cm/sec;p=0.01], higher E/e’av [9.3(3.7-13.61) vs 6.7(4.5-11.2);p=0.0001], increased SRV [17.1(11.9-28) vs 15.3(12.2-22.1) cm/sec;p=0.018], higher CI [3.3(2.1-4.7) vs 2.9(2.1-5.1) L/min/m2;p=0.001], increased PASP [35(30-43) vs 30(18-45) mmHg;p=0.003], increased TRV [2.7(2.3-3) vs 2.5(1.8-3) m/sec;p=0.028], decreased SVRI [1811(1355-3580) vs 2527.5(1396-3607) dyn/s/m^2^/cm-^5^;p=0.0001], increased E(s) [83(50-182) vs 74(49-116) cm/sec;p=0.0001], higher E/e’av(s) [8(3.2-10.23) vs 6.1(3.8-7.9);p=0.003] and increased TAPSE(s) [29(23-34) vs 28(21-34) mm;p=0.021], compared to patients with Child-Pugh stage A respectively. Table 3 highlights the differences of the comparing variables between the two groups.

**Table 3.** Comparison of patients according to the Child-Pugh stage

| Parameter | Child-Pugh A | Child-Pugh B/C | p value |
| --- | --- | --- | --- |
| Age (years) | 57 (33-78) | 57.5 (38-77) | NS |
| HR (bpm) | 77 (59-110) | 78.5 (59-91) | NS |
| QTc (msec) | 428 (368-490) | 454 (416-497) | 0.004 |
| EF (%) | 65 (56-75) | 65 (57-70) | NS |
| E (cm/sec) | 72 (53-119) | 90 (51-128) | 0.032 |
| BNP (pg/ml) | 18.5 (5-145) | 46 (6.8-369) | 0.016 |
| A (cm/sec) | 72 (42-120) | 87 (53-127) | 0.001 |
| E/A (ratio) | 1 (0.6-1.9) | 0.9 (0.56-1.85) | NS |
| DT (msec) | 219 (121-262) | 227.5 (169-272) | NS |
| IVRT (msec) | 111 (83-130) | 100.5 (70-127) | NS |
| TAPSE (mm) | 24 (21-29) | 25 (21-37) | NS |
| e' septal (cm/sec) | 8.9 (5.3-15.3) | 8.4 (5-14.2) | NS |
| e' lateral (cm/sec) | 12.2 (8.8-19.7) | 11.5 (7-17) | NS |
| E/e' av (ratio) | 6.7 (4.5-11.2) | 9.3 (3.7-13.6) | 0.0001 |
| SRV (cm/sec) | 15.3 (12.2-22.1) | 17.1 (11.9-28) | 0.018 |
| CI (L/min/m<sup>2</sup>) | 2.9 (2.1-5.1) | 3.3 (2.1-4.7) | 0.0001 |
| LA vol (ml/m<sup>2</sup>) | 23.3 (12.1-42.9) | 24.4 (18.1-39.3) | NS |
| LVEDD (mm) | 49.5 (40-62) | 51 (42-62) | NS |
| LVESD (mm) | 31 (22-37) | 31 (24-37) | NS |
| PASP (mmHg) | 30 (18-45) | 35 (30-43) | 0.003 |
| TRV (m/sec) | 2.5 (1.8-3) | 2.7 (2.3-3) | 0.028 |
| SVRI (dyn/s/m<sup>2</sup>/cm<sup>-5</sup>) | 2527.5 (1396-3607) | 1811 (1355-3580) | 0.0001 |
| E(s) (cm/sec) | 74 (49-116) | 83 (50-182) | 0.0001 |
| e'septal(s) (cm/sec) | 10.2 (6.8-18.8) | 10.5 (5.5-20.5) | NS |
| e'lateral(s) (cm/sec) | 13.2 (10-18.4) | 13.9 (7.3-22) | NS |
| E/e'av(s) (ratio) | 6.1 (3.8-7.9) | 8 (3.2-10.2) | 0.003 |
| TAPSE(s) (mm) | 28 (21-34) | 29 (23-34) | 0.021 |
| SRV(s) (cm/sec) | 24 (12.3-41) | 26.3 (13.3-41) | NS |
| CI(s) (L/min/m<sup>2</sup>) | 5.1 (2.4-8) | 5.3 (3.1-10.5) | NS |
| ΔSRV (cm/sec) | 0.53 (-0.07 – 1.8) | 0.43 (0.001-1.32) | NS |
| ΔTAPSE (mm) | 0.13 (-0.18 – 0.42) | 0.12 (-0.14 – 0.33) | NS |
| ΔCI (L/min/m<sup>2</sup>) | 0.8 (-0.04 – 2.5) | 0.66 (0.13-1.76) | NS |
| ΔE/e'av | -0.07 (-0.44 – 0.3) | -0.15 (-0.43 – 0.06) | NS |

Variables are expressed as median (range) values
HR: heart rate; LVEDD: left ventricular end diastolic diameter; LVESD: left ventricular end systolic diameter; LA vol: left atrium volume; EF: ejection fraction at rest; CI: cardiac index; SVRI: systemic vascular resistance index; E: peak early filling velocity during early ventricle diastole; A: late diastolic filling velocity during atrial systole; DT: deceleration time of E wave; IVRT: isovolumetric relaxation time; e'septal: early diastolic mitral annular velocity from the septal side; e'lateral: early diastolic mitral annular velocity from the lateral side; e'av: average early diastolic mitral annular velocity; PASP: pulmonary artery systolic pressure; TRV: tricuspid regurgitation velocity; TAPSE: tricuspid annular plane systolic excursion; SRV: systolic right ventricular function; Cis(s) CI after stress; e'septal(s): early diastolic mitral annular velocity from the septal side after stress; TAPSE(s): TAPSE after stress; SRV(s): SRV after stress; TRV(s): TRV after stress.

### Correlations between Child-Pugh score and sonographic parameters

A statistically significant positive correlation was verified between Child-Pugh score and QTc (r=0.356, p=0.001), E (r=0.29, p=0.006), BNP (r=0.347, p=0.001), A (r=0.379, p=0.0001), E/e’av (r=0.418, p=0.0001), CI (r=0.54, p-0.0001), PASP (r=0.278, p=0.009), E(s) (r=0.418, p=0.0001), E/e’av(s) (r=0.321, p=0.002) and TAPSE(s) (r=0.291, p=0.01), while a trend forward a positive correlation with TRV was reported (r=0.209, p=0.051). A significant negative relationship between Child-Pugh score and SVRI was revealed (r=-0.595, p=0.0001).

## Discussion

In cirrhosis a hyperdynamic circulation is developed along with the aggravation of liver dysfunction and portal hypertension. Furthermore, in some patients a blunted cardiac function defined as cirrhotic cardiomyopathy (CC) is also present, enhancing further the circulatory dysfunction of cirrhosis [7]. The prevalence of this entity remains controversial, as several studies have shown conflicting results [14-16]. Different criteria selected for the evaluation of CC at each of these studies, could probably explain this argument. According to Montreal criteria, the prevalence of LVDD which is the usual primary component of CC, has been described up to 70% [7]. In a previous study from our group, using the 2009 criteria of the American Society of Echocardiography, a prevalence close to 37% was demonstrated [15]. In the current study, the percentage of LVDD was estimated about 80%, 40% and 11%, when the Montreal criteria, the 2009 criteria, or the 2016 guidelines of the American Society of Echocardiography and the European Association of Cardiovascular Imaging, modified in 2019 by the CC consortium respectively applied. Razpotnik et al had also illustrated this correspondence between the percentages of patients identified with LVDD and the criteria that had been used for this evaluation. Thus, according to Montreal criteria the authors showed a prevalence of 67%, which dropped to 35% and 7.5% by applying the 2009 and 2019 criteria respectively [26].

We denoted a “good” agreement between the Montreal and the 2009 guidelines (k value: 0.754). By contrast, the recent guidelines had not pointed such a strong correlation with the aforementioned, as the agreement with each of them was “fair” (k value: 0.338 and 0.316 for comparison to Montreal and 2016 criteria respectively). Based on the 2019 guidelines, the development of LVDD in our cohort was not found to significantly correlate with gender, etiology of liver disease (alcoholic vs non-alcoholic), or Child-Pugh stage. These results are in accordance with those formerly presented by our group [15].

The 2019 algorithm is more complicated, but probably more appropriate for the estimation of LVDD in cirrhotic patients, as it combines several factors, less dependable on the alterations of preload and afterload which are being affected in cirrhosis [18]. Nevertheless, it seems that regardless of its good specificity, its sensitivity and negative predictive value (NPV) are moderate. Obokata et al have reported sensitivity rates of 34% and NPV of 53% in patients with diastolic dysfunction and preserved EF%. The addition of stress test has been considered to increase the sensitivity and the NPV [27]. Therefore, in order to detect any latent LVDD not present at rest, we re-evaluated the diastolic cardiac function after the administration of dobutamine. Notably, not even one patient without LVDD at rest, accomplished the LVDD criteria during the stress test.

Patients with LVDD compared to those without, had no differences regarding the Child-Pugh score and the MELD score, corroborating that the presence of LVDD is not associated with the severity of liver cirrhosis. However, patients with LVDD had a significant longer QTc, a trend for higher BNP and a trend to older age. QTc interval prolongation has been correlated with the severity and complications of cirrhosis in several previous studies, but data are still controversial [28,29]. In our study, a significant but not so strong correlation was found between the QTc and Child-Pugh score. Considering BNP, it has been suggested that high levels are implicated with increased post-transplant mortality [30].

Several studies have shown significant correlation between E/e’av and filling cardiac pressures [31,32]. Ommen et al verified the excellent specificity of E/e’av although they identified many patients with increased filling pressures despite of a normal E/e’av, arising questions about its sensitivity. They concluded that an increased E/e’av strongly supports the existence of high left filling pressures and high pulmonary capillary wedge pressure and thus LVDD, but a normal E/e’av does not exclude LVDD [31]. In our study, the limited predictability of E/e’av in differentiating LVDD was confirmed, as it did not significantly differ between patients with and without LVDD.

Interestingly, neither LVSD, nor RVSD, were identified at rest. A minority of patients (4.5%) was unable to increase adequately the CI (ΔCI<10%) during stress, which is an indication of LVSD. On the contrary, no RVSD was verified even during stress. However, the significantly lower TAPSE(s), SRV(s) and ΔSRV after stress in patients with LVDD, points to a tendency towards right systolic derangement.

When patients with Child-Pugh stage A were compared to those with B/C, we reported that the latter group had significantly lower SVRI and higher CI. This finding was expected due to the more advanced splanchnic vasodilation and hyperdynamic circulation at the later stages of cirrhosis. Furthermore, this group had longer QTc, higher BNP, increased PASP, increased SRV, higher E/e’av and a trend towards increased TAPSE, making clear that several parameters related with the severity of LVDD are significantly exacerbated in this advanced stage of cirrhosis. This finding was further supported by the significant positive correlation found between the E/e’av and the Child-Pugh score. Moreover, the significantly increased E/e’av(s) and TAPSE(s) after stress in Child-Pugh B/C patients, combined with the significant positive correlation found between the E/e’av(s) or TAPSE(s) and the Child-Pugh score respectively, are indicative of further cardiac deterioration during stress in patients with advanced liver disease. These findings point out the concerns about the potential risk for clinical cardiac derangement in advanced cirrhotic patients during a stressful invasive procedure, such as TIPS implementation or liver transplantation [33-35].

Our study has some limitations. First, the non-existence of a control group. However, our purpose was not to compare the prevalence of cardiac dysfunction between cirrhotic and non-cirrhotic patients, but to determine any differences in prevalence of CC in cirrhotic patients according to the different diagnostic algorithms that have been proposed. Secondly, the systolic dysfunction was not evaluated by measuring the global longitudinal strain which seems capable of identifying abnormal contraction patterns in the setting of apparently normal EF% [36]. Albeit, this method is not included in the latest guidelines. As an alternative we performed a dobutamine stress test. The ability of the latter in detecting abnormalities, mainly changes in volumes and EF% measured by TDI, is still a matter of debate [37-39]. Cardiovascular magnetic resonance using dobutamine stress is considered a superior method [40]. Unfortunately, this is not available in our center. Nevertheless, the low sensitivity of TDI dobutamine stress seems to be attributed to the delayed response of cirrhotic patients to inotropic stimuli and therefore, higher doses of dobutamine or an extension in time are required [40]. In our study, we administrated the maximum dobutamine dose in the majority of our patients and if possible, we extended test’s duration until the achievement of the maximum response.

The strength of our study is the evaluation of diastolic and systolic function of both ventricles at rest and after stress. Of note, it is the first time that such a comprehensive approach has been initiated.

In conclusion, prevalence of CC is lower when estimated according to the more recent guidelines. LVSD is usually absent at rest, while a small percentage of cases is recognized during stress. Interestingly, RVSD is not present neither at rest, nor at stress, but some ultrasonographic parameters related to right systolic contractility, aggravate during stress. The predominant component of CC is LVDD, whose presence is independent of the stage of cirrhosis, but its severity correlates with the degree of liver dysfunction. The exacerbation of several diastolic and systolic parameters during stress, particularly in decompensated cirrhotic patients, raises questions about the potential risk of this group of patients to progress to clinical heart failure during any invasive intervention, such as TIPS insertion, or liver transplantation.

## Data Availability

All data produced in the present work are contained in the manuscript

## Abbreviations

CC: cirrhotic cardiomyopathy
EF: ejection fraction
TIPS: transjugular intrahepatic portosystemic shunt
LVDD: left ventricular diastolic dysfunction
2D-SWE: two-dimension shear wave elastography
ECG: electrocardiograph
QTc: corrected QTc
PW: pulsed wave
TDI: tissue doppler imaging
HR: heart rate
LVEDD: left ventricular end diastolic diameter
LVESD: left ventricular end systolic diameter
LA vol: left atrium volume
EF: ejection fraction at rest
CI: cardiac index
SVRI: systemic vascular resistance index
E: peak early filling velocity during early ventricle diastole
A: late diastolic filling velocity during atrial systole
DT: deceleration time of E wave
IVRT: isovolumetric relaxation time
e’septal: early diastolic mitral annular velocity from the septal side
e’lateral: early diastolic mitral annular velocity from the lateral side
e’av: average early diastolic mitral annular velocity
PASP: pulmonary artery systolic pressure
TRV: tricuspid regurgitation velocity
TAPSE: tricuspid annular plane systolic excursion
SRV: systolic right ventricular function
CI(s): CI after stress
e’septal(s): early diastolic mitral annular velocity from the septal side after stress
e’lateral(s): early diastolic mitral annular velocity from the lateral side after stress
TAPSE(s): TAPSE after stress
SRV(s): SRV after stress
TRV(s): TRV after stress
BNP: brain natriuretic peptide
RVSD: right ventricular systolic dysfunction
NPV: negative predictive value.

## References

1. Iwakiri Y, Groszmann RJ. The hyperdynamic circulation of chronic liver diseases: from the patient to the molecule. Hepatology 2006;43:S121–S131.

2. Wiest R, Garcia-Tsao G. Bacterial translocation (BT) in cirrhosis. Hepatology 2005;41:422–433.

3. Vlachogiannakos J, Saveriadis AS, Viazis N, et al. Intestinal decontamination improves liver haemodynamics in patients with alcohol-related decompensated cirrhosis. Aliment Pharmacol Ther. 2009;29:992–999.

4. Moller S, Henriksen JH. Cirrhotic cardiomyopathy. J Hepatol 2010;53:179–190.

5. Alqahtani SA, Fouad TR, Lee SS. Cirrhotic cardiomyopathy. Semin Liver Dis 2008;28:59–69.

6. Zambruni A, Trevisani F, Caraceni P, Bernardi M. Cardiac electrophysiological abnormalities in patients with cirrhosis. J Hepatol 2006;44:994–1002.

7. Karagiannakis DS, Papatheothoridis G, Vlachogiannakos J. Recent advances in cirrhotic cardiomyopathy. Dig Dis Sci 2015;60:1141–1151.

8. Venon WD, Pumo SL, Imperatrice B, et al. Transjugular intrahepatic portosystemic shunt in refractory ascites: clinical impact of left ventricular diastolic dysfunction. Eur J Gastroenterol Hepatol 2021;33(Suppl. 1):e464–e470.

9. Carvalheiro F, Rodrigues C, Adrego T, et al. Diastolic Dysfunction in Liver Cirrhosis: Prognostic Predictor in Liver Transplantation? Transplant Proc 2016;48:128–31.

10. Ruiz-del-Arbol L, Achecar L, Serradilla R, et al. Diastolic dysfunction is a predictor of poor outcomes in patients with cirrhosis, portal hypertension and normal creatinine. Hepatology 2013;58:1732–1741.

11. Karagiannakis DS, Vlachogiannakos J, Anastasiadis G, et al. Diastolic cardiac dysfunction is a predictor of dismal prognosis in patients with liver cirrhosis. Hepatol Int 2014;8:588–594.

12. Alexopoulou A, Papatheodoridis G, Pouriki S, et al. Diastolic myocardial dysfunction does not affect survival in patients with cirrhosis. Transpl Int 2012;25:1174–1181.

13. Merli M, Torromeo C, Giusto M, et al. Survival at 2 years among liver cirrhotic patients is influenced by left atrial volume and left ventricular mass. Liver Int 2017;37:700–706.

14. Torregrosa M, Aguadé S, Dos L, et al. Cardiac alterations in cirrhosis: reversibility after liver transplantation. J Hepatol 2005;42:68–74.

15. Dimitrios S. Karagiannakis DS, Vlachogiannakos J, Anastasiadis G, et al. Frequency and severity of cirrhotic cardiomyopathy and its possible relationship with bacterial endotoxemia. Dig Dis Sci 2013;58:3029–36.

16. Almeida JG, Fontes-Carvalho R, Sampaio F, et al. Impact of the 2016 ASE/EACVI recommendations on the prevalence of diastolic dysfunction in the general population. Eur Heart J Cardiovasc Imaging 2018;19:380–6.

17. Nagueh SF, Smiseth OA, Appleton CP, et al. Recommendations for the evaluation of left ventricular diastolic function by echocardiography: an update from the American Society of Echocardiography and the European Association of Cardiovascular Imaging. J Am Soc Echocardiogr 2016;29:277–314.

18. Izzy M, Van Wagner LB, Lin G, et al. Redefining cirrhotic cardiomyopathy for the modern era. Hepatology 2019;0:1–12.

19. Barr RG, Wilson SR, Rubens D, et al. Update to the Society of Radiologists in Ultrasound Liver Elastography Consensus Statement. Radiology 2020;296:263–274.

20. Guilo P, Mansourati J, Maheu B, et al. Long-term prognosis in patients with alcoholic cardiomyopathy and severe heart failure after total abstinence. Am J Cardiol 1997;79:1276–1278.

21. Cerqueira MD, Weissman NJ, Dilsizian V, et al. Standardized myocardial segmentation and nomenclature for tomographic imaging of the heart, A statement for healthcare professionals from the Cardiac Imaging Committee of the Council on Clinical Cardiology of the American Heart Association. Circulation 2002;105:539–42.

22. Paulus WJ, Tschöpe C, Sanderson JE, et al. How to diagnose diastolic heart failure: a consensus statement on the diagnosis of heart failure with normal left ventricular ejection fraction by the Heart Failure and Echocardiography Associations of the European Society of Cardiology. Eur Heart J 2007;28:2539–2550.

23. Nagueh SF, Appleton CP, Gillebert TC, et al. Recommendations for the evaluation of left ventricular diastolic function by echocardiography. J Am Soc Echocardiogr 2009;22:107–133.

24. Modin D, Møgelvang R, Andersen DM, et al. Right Ventricular Function evaluated by Tricuspid Annular Plane Systolic Excursion predicts cardiovascular death in the general population. J Am Heart Assoc 2019;8:e012197.

25. Kharbanda RK, Moore JP, Yannick J H J Taverne YJH et al. Cardiac resynchronization therapy for the failing systemic right ventricle: A systematic review. Int J Cardiol 2020;318:74–81.

26. Razpotnik M, Bota S, Wimmer P, et al. The prevalence of cirrhotic cardiomyopathy according to different diagnostic criteria. Liver Int 2021;41:1058–1069.

27. Obokata M, Kane GC, Reddy YNV, et al. The role of diastolic stress testing in the evaluation for HFpEF: A simultaneous invasive-echocardiographic study. Circulation 2017;135: 825–838.

28. Koshy AN, Gow PJ, Testro A, et al. Relationship between QT interval prolongation and structural abnormalities in cirrhotic cardiomyopathy: A change in the current paradigm. Am J Transplant 2021;21:2240–2245.

29. Li S, Hao X, Liu S, et al. Prolonged QTc interval predicts long-term mortality in cirrhosis: a propensity score matching analysis. Scand J Gastroenterol 2021;56:570–577.

30. Kwon H-M, Moon Y-J, Kim K-S, et al. Prognostic value of B-Type Natriuretic Peptide in liver transplant patients: Implication in post-transplant mortality. Hepatology 2021;74:336–350.

31. Ommen SR, Nishimura RA, Appleton CP, et al. Clinical utility of doppler echocardiography and tissue doppler imaging in the estimation of left ventricular filling pressures: A comparative simultaneous doppler-catheterization study. Circulation 2000;102:1788–1794.

32. Nagueh SF, Middleton KJ, Kopelen HA, et al. Doppler tissue imaging: A noninvasive technique for evaluation of left ventricular relaxation and estimation of filling pressures. J Am Coll Cardiol 1997;30:1527–1533.

33. Billey C, Billet S, Robic MA, et al. A prospective study identifying predictive factors of cardiac decompensation after Transjugular Intrahepatic Portosystemic Shunt: The Toulouse Algorithm. Hepatology 2019;70:1928–1941.

34. Mester RA, Stoll WD. Cirrhotic Cardiomyopathy and cardiac failure after liver transplantation: A case series. Transplant Proc 2021;53:2354–2357.

35. Liu H, Jayakumar S, Traboulsi M, et al. Cirrhotic cardiomyopathy: Implications for liver transplantation. Liver Transpl 2017;23:826–835.

36. Cimino S, Canali E, Petronilli V, et al. Global and regional longitudinal strain assessed by two-dimensional speckle tracking echocardiography identifies early myocardial dysfunction and transmural extent of myocardial scar in patients with acute ST elevation myocardial infarction and relatively preserved LV function. Eur Heart J Cardiovasc Imaging 2013;14:805–811.

37. Kim MY, Baik SK, Won CS, et al. Dobutamine stress echocardiography for evaluating cirrhotic cardiomyopathy in liver cirrhosis. Korean J Hepatol. 2010;16:376–82.

38. Dahl EK, Møller S, Kjær A, et al. Diastolic and autonomic dysfunction in early cirrhosis: a dobutamine stress study. Scand J Gastroenterol 2014;49:362–72.

39. Krag A, Bendtsen F, Dahl EK, et al. Cardiac function in patients with early cirrhosis during maximal beta-adrenergic drive: a dobutamine stress study. PLoS One 2014;9:e109179.

40. Sampaio F, Lamata P, Bettencourt N, et al. Assessment of cardiovascular physiology using dobutamine stress cardiovascular magnetic resonance reveals impaired contractile reserve in patients with cirrhotic cardiomyopathy. J Cardiovasc Magn Reson 2015;17:61.

